# Racial and Ethnic Differences in Cesarean Delivery Across Insurance Types, United States, 2014–2024

**DOI:** 10.64898/2026.04.04.26350151

**Authors:** Oluwasegun Akinyemi, Mojisola Fasokun, Delia Singleton, Fadeke Ogunyankin, Nicholas Mercado, Samar Khalil, Kaelyn Gordon, Miriam Micheal, Kakra Hughes, Guoyang Luo, Shari Lawson, Eke Ahizechukwu

**Author notes:** Corresponding Author: Delia Singleton, 2041 Georgia Ave NW #4002, Washington, DC 20060, USA, Race–Insurance differences in Cesarean Delivery.

## Abstract

**Introduction:** Cesarean delivery accounts for nearly one-third of U.S. births and is associated with substantial maternal morbidity and health care costs. Persistent racial disparities have been documented, yet the structural factors contributing to these differences remain incompletely understood. The extent to which insurance coverage shapes racial disparities in cesarean delivery remains unclear.

**Objective:** To evaluate the independent and interactive associations of race/ethnicity and insurance coverage with cesarean delivery in the United States.

**Methods:** Population-based retrospective cohort study using singleton live births recorded in the United States Vital Statistics Natality files from 2014 to 2024. Multivariable logistic regression was used to estimate the independent effects of race/ethnicity and insurance status on cesarean delivery, including interaction terms to test effect modification, using national birth certificate data. Models were adjusted for maternal demographics, clinical factors, and temporal covariates. Adjusted odds ratios, predicted probabilities, and absolute risk differences were derived from post-estimation marginal effects. The main outcome measure was cesarean delivery (yes vs no).

**Results:** Among 41,543,568 deliveries from 2014 to 2024, 13,312,221 (32.0%) were cesarean deliveries. After adjustment, both race and ethnicity and insurance status were independently associated with cesarean delivery. Compared with non-Hispanic White women, non-Hispanic Black women had higher odds of cesarean delivery (odds ratio [OR], 1.22; 95% CI, 1.22–1.23). Relative to uninsured women, those with private insurance had 59% higher odds of cesarean delivery (OR, 1.59; 95% CI, 1.58–1.60). Significant interaction effects were observed, indicating that insurance coverage modified racial and ethnic differences in cesarean delivery. Non-Hispanic Black women had the highest predicted probabilities across all insurance categories, with the largest absolute disparities observed among uninsured women.

**Conclusion:** Racial and ethnic differences in cesarean delivery persist in the United States and are modified by insurance coverage, suggesting that coverage-related differences may contribute to inequities in obstetric care.

## Introduction

Cesarean delivery is the most frequently performed surgical procedure in the United States, accounting for nearly one-third of all births [1–3]. While cesarean delivery is vital for many high-risk pregnancies, overuse without clear clinical indications increases maternal morbidity, prolongs recovery, raises healthcare costs, and heightens risks in subsequent pregnancies [4–7]. Reducing unnecessary cesarean delivery while ensuring equitable access to medically indicated care remains a major public health and obstetric priority.

Racial and ethnic disparities in cesarean delivery are well documented in the United States [8–11]. Non-Hispanic Black women consistently experience higher cesarean delivery rates than Non-Hispanic White women, even after adjustment for obstetric risk factors and comorbidities [8,9,11–13]. In contrast, Hispanic, Asian, and American Indian or Alaska Native women often have lower or more variable cesarean rates [11,14]. These persistent differences suggest that non-clinical factors including structural racism, differential access to high-quality obstetric care, provider decision-making, and institutional practices play a substantial role in shaping delivery mode [8,9,15,16].

Insurance coverage is another key determinant of obstetric care, shaping both access and delivery practices [17–20]. Privately insured women generally have higher cesarean rates than Medicaid beneficiaries, whereas uninsured women tend to have lower rates, potentially reflecting differences in reimbursement incentives, access to services, and patterns of care delivery [21–24]. Importantly, the effect of insurance status may not be uniform across racial and ethnic groups [23]. Instead, insurance coverage may modify existing disparities, either exacerbating or attenuating inequities in cesarean delivery [22].

Despite extensive research on racial disparities and insurance-based differences in cesarean delivery, few contemporary national studies have explicitly examined the interaction between race/ethnicity and insurance status [8,23]. Moreover, many prior studies rely on older datasets or focus solely on relative measures, limiting their relevance for current policy and quality improvement efforts [10,18,25].

Using United States Vital Statistics Natality data from 2014 to 2024 [26,27], this study aimed to assess the independent associations of race/ethnicity and insurance status with cesarean delivery and to determine whether insurance coverage modifies racial and ethnic disparities in cesarean delivery. By applying interaction-based analyses to a decade of population-level data, this study provides updated evidence on how race, ethnicity, and insurance intersect to influence cesarean delivery patterns in the United States.

## Methods

### Study Design and Data Source

We conducted a population-based retrospective cohort study using the United States Vital Statistics Natality files from January 1, 2014, through December 31, 2024 [26]. These files capture all live births occurring in the United States and include standardized information on maternal sociodemographic characteristics, clinical conditions, obstetric history, and delivery outcomes [28]. Analyses adhered to the STROBE reporting guidelines for observational studies. The study used de-identified NCDB data and was deemed exempt from institutional review board oversight; informed consent was not required [29].

### Study Population

The analytic sample included all singleton live births during the study period. We excluded multifetal gestations and records with missing information on mode of delivery, race/ethnicity, or insurance status. These exclusions were made a priori because multifetal pregnancies have distinct clinical indications for cesarean delivery and incomplete records preclude valid estimation of exposure–outcome associations.

### Exposure Variables

The primary exposure was maternal race and ethnicity, categorized as Non-Hispanic White (reference), Non-Hispanic Black, Hispanic (any race), Non-Hispanic Asian, Non-Hispanic American Indian or Alaska Native, Non-Hispanic Native Hawaiian or Other Pacific Islander, Non-Hispanic Multiracial, and Unknown/not stated. The secondary exposure was insurance status at delivery, classified as Medicaid (reference), private insurance, uninsured (self-pay), other insurance, or unknown.

### Outcome Measures

The primary outcome was cesarean delivery, defined as delivery by cesarean section versus vaginal delivery.

### Covariates

Covariates were selected a priori based on clinical relevance and prior literature and included maternal age, marital status, educational attainment, parity, prior cesarean delivery, pre-pregnancy hypertension, gestational hypertension or preeclampsia, diabetes, smoking during pregnancy, prenatal care utilization, and year of delivery.

### Statistical Analysis

We used multivariable logistic regression to estimate adjusted odds ratios (ORs) and 95% confidence intervals (CIs) for the independent associations of race/ethnicity and insurance status with cesarean delivery. Effect modification was assessed by including interaction terms between race/ethnicity and insurance status. Adjusted predicted probabilities and absolute risk differences were calculated using post-estimation marginal effects derived via the delta method. Robust standard errors were used, and statistical significance was defined as a two-sided P value <0.05. Analyses were conducted using Stata.

### Sensitivity Analyses

Sensitivity analyses included re-estimating models excluding women with a prior cesarean delivery and restricting analyses to term births to assess the robustness of findings.

### Ethical Considerations

The NCHS natality files are de-identified and publicly available. Consequently, this study is considered non-human subjects research and did not require institutional review board approval or informed consent.

## Result

### Cohort and Baseline Characteristics

Among 41,543,568 births, 13,312,221 (32.04%) were delivered by cesarean section and 28,231,347 (67.96%) by vaginal delivery. Individuals undergoing cesarean delivery were older (mean [SD], 30.20 [5.83] vs 28.54 [5.76] years; P<.001) and were more likely to be non-Hispanic Black (16.03% vs 13.25%) and privately insured (51.65% vs 48.69%) compared with those delivering vaginally (all P<.001). Medicaid remained the most common payer overall (41.70%) (Table 1).

**Table 1.** Baseline Maternal and Clinical Characteristics by Mode of Delivery Among U.S. Births, 2014–2024. Data are presented as number (percentage) unless otherwise indicated. Percentages may not sum to 100 due to rounding. AIAN indicates American Indian or Alaska Native; NHOPI, Native Hawaiian or Other Pacific Islander; GED, General Educational Development; SES, socioeconomic status. Chi-square tests were used to compare categorical variables, and analysis of variance was used for continuous variables. ᵃ F-statistic from one-way ANOVA; all other statistics are Pearson chi-square. All P values are two-sided.

| Variables | Total<br>(N=41,543,568) | Cesarean<br>Delivery<br>(n=13,312,221;<br>32.04%) | Vaginal Delivery<br>(n=28,231,347;<br>67.96%) | Chi-square | P-value |
| --- | --- | --- | --- | --- | --- |
| **Age (years), mean ± SD** | 29.07 ± 5.83 | 30.20 ± 5.83 | 28.54 ± 5.76 | 7.4×10<sup>5a</sup> | <0.001 |
| **Race/Ethnicity** |  |  |  | 6.6×10<sup>4</sup> | <0.001 |
| Non-Hispanic White | 21,168,383<br>(51.20) | 6,556,992 (49.52) | 14,611,391 (51.99) |  |  |
| Non-Hispanic Black | 5,846,571<br>(14.14) | 2,122,592 (16.03) | 3,723,979 (13.25) |  |  |
| Non-Hispanic AIAN | 310,906 (0.75) | 89,439 (0.68) | 221,467 (0.79) |  |  |
| Non-Hispanic Asian | 2,551,426 (6.17) | 850,805 (6.43) | 1,700,621 (6.05) |  |  |
| Non-Hispanic NHOPI | 106,056 (0.26) | 33,559 (0.25) | 72,497 (0.26) |  |  |
| Non-Hispanic Multiracial | 924,721 (2.24) | 278,876 (2.11) | 645,845 (2.30) |  |  |
| Hispanic (any race) | 10,066,605<br>(24.35) | 3,192,418 (24.11) | 6,874,187 (24.46) |  |  |
| Unknown/not stated | 369,767 (0.89) | 115,504 (0.87) | 254,263 (0.90) |  |  |
| **Insurance Type** |  |  |  | 8.9×10<sup>4</sup> | <0.001 |
| Medicaid | 17,237,757<br>(41.70) | 5,464,441 (41.28) | 11,773,316 (41.90) |  |  |
| Private | 20,517,846<br>(49.64) | 6,836,734 (51.65) | 13,681,112 (48.69) |  |  |
| Uninsured (self-pay) | 1,767,037 (4.28) | 407,712 (3.08) | 1,359,325 (4.84) |  |  |
| Other | 1,516,447 (3.67) | 445,771 (3.37) | 1,070,676 (3.81) |  |  |
| Unknown/not stated | 294,020 (0.71) | 82,001 (0.62) | 212,019 (0.75) |  |  |
| **Education Level** | | | | $4.9 \times 10^4$ | <0.001 |
| < High school | 5,141,364<br>(12.62) | 1,459,374 (11.19) | 3,681,990 (13.30) |  |  |
| High school/GED | 10,587,574<br>(25.99) | 3,303,248 (25.32) | 7,284,326 (26.30) |  |  |
| Some college/Associate | 11,310,668<br>(27.76) | 3,756,924 (28.80) | 7,553,744 (27.28) |  |  |
| Bachelor's or higher | 13,698,297<br>(33.63) | 4,526,479 (34.70) | 9,171,818 (33.12) |  |  |
| **Marital Status<sup>b</sup>** | | | | $3.4 \times 10^3$ | <0.001 |
| Unmarried | 6,140,837<br>(40.07) | 1,916,411 (39.01) | 4,224,426 (40.58) |  |  |
| Married | 9,183,023<br>(59.93) | 2,996,327 (60.99) | 6,186,696 (59.42) |  |  |
| **Gestational Age at Delivery** | | | | $8.6 \times 10^5$ | <0.001 |
| Term ($\geq 37$ weeks) | 37,337,289<br>(89.93) | 11,127,612 (83.61) | 26,209,677 (92.90) | | |
| Preterm (<37 weeks) | 4,182,879<br>(10.07) | 2,181,206 (16.39) | 2,001,673 (7.10) |  |  |
| **Previous Preterm Birth** | | | | $4.9 \times 10^4$ | <0.001 |
| No | 39,846,278<br>(96.40) | 12,645,574 (95.53) | 27,200,704 (96.81) |  |  |
| Yes | 1,443,792 (3.49) | 581,607 (4.39) | 862,185 (3.07) |  |  |
| Unknown | 43,037 (0.10) | 9,478 (0.07) | 33,559 (0.12) |  |  |
| **Pre-pregnancy Diabetes** | | | | $1.4 \times 10^5$ | <0.001 |
| No | 40,873,661<br>(98.89) | 12,980,728 (98.07) | 27,892,933 (99.28) |  |  |
| Yes | 416,409 (1.01) | 246,453 (1.86) | 169,956 (0.60) |  |  |
| Unknown | 43,037 (0.10) | 9,478 (0.07) | 33,559 (0.12) |  |  |
| **Gestational Diabetes** | | | | $1.5 \times 10^5$ | <0.001 |
| No | 38,378,303<br>(92.95) | 12,000,363 (90.73) | 26,377,940 (94.00) |  |  |
| Yes | 2,911,767 (7.05) | 1,226,818 (9.27) | 1,684,949 (6.00) |  |  |
| **Pre-pregnancy Hypertension** | | | | $1.6 \times 10^5$ | <0.001 |
| No | 40,329,470<br>(97.67) | 12,740,617 (96.32) | 27,588,853 (98.31) |  |  |
| Yes | 960,600 (2.33) | 486,564 (3.68) | 474,036 (1.69) |  |  |
| **Gestational Hypertension** | | | | $1.9 \times 10^5$ | <0.001 |
| No | 38,096,034<br>(92.26) | 11,851,263 (89.60) | 26,244,771 (93.52) |  |  |
| Yes | 3,194,036 (7.74) | 1,375,918 (10.40) | 1,818,118 (6.48) |  |  |
| **Previous Cesarean Delivery** |  |  |  | 1.0×10<sup>7</sup> | <0.001 |
| No | 34,945,113<br>(84.63) | 7,738,537 (58.50) | 27,206,576 (96.95) |  |  |
| Yes | 6,344,957<br>(15.37) | 5,488,644 (41.50) | 856,313 (3.05) |  |  |
| **SES Disadvantage Score (0-2)** |  |  |  | 1.8×10<sup>4</sup> | <0.001 |
| 0 (Least disadvantaged) | 19,282,084<br>(46.41) | 6,362,077 (47.79) | 12,920,007 (45.76) |  |  |
| 1 | 11,556,273<br>(27.82) | 3,673,225 (27.59) | 7,883,048 (27.92) |  |  |
| 2 (Most disadvantaged) | 10,705,211<br>(25.77) | 3,276,919 (24.62) | 7,428,292 (26.31) |  |  |
| **Birth Year** |  |  |  | 859.1 | <0.001 |
| 2014 | 3,994,520 (9.62) | 1,287,900 (9.67) | 2,706,620 (9.59) |  |  |
| 2015 | 3,986,186 (9.60) | 1,275,980 (9.59) | 2,710,206 (9.60) |  |  |
| 2016 | 3,953,472 (9.52) | 1,261,949 (9.48) | 2,691,523 (9.53) |  |  |
| 2017 | 3,862,502 (9.30) | 1,235,362 (9.28) | 2,627,140 (9.31) |  |  |
| 2018 | 3,799,906 (9.15) | 1,211,372 (9.10) | 2,588,534 (9.17) |  |  |
| 2019 | 3,755,247 (9.04) | 1,189,718 (8.94) | 2,565,529 (9.09) |  |  |
| 2020 | 3,617,673 (8.71) | 1,150,672 (8.64) | 2,467,001 (8.74) |  |  |
| 2021 | 3,666,952 (8.83) | 1,176,445 (8.84) | 2,490,507 (8.82) |  |  |
| 2022 | 3,673,216 (8.84) | 1,181,015 (8.87) | 2,492,201 (8.83) |  |  |
| 2023 | 3,602,345 (8.67) | 1,165,226 (8.75) | 2,437,119 (8.63) |  |  |
| 2024 | 3,631,549 (8.74) | 1,176,582 (8.84) | 2,454,967 (8.70) |  |  |

Data are presented as number (percentage) unless otherwise indicated. Percentages may not sum to 100 due to rounding. AIAN indicates American Indian or Alaska Native; NHOPI, Native Hawaiian or Other Pacific Islander; GED, General Educational Development; SES, socioeconomic status. Chi-square tests were used to compare categorical variables, and analysis of variance was used for continuous variables. <sup>a</sup> F-statistic from one-way ANOVA; all other statistics are Pearson chi-square. All P values are two-sided.

Cesarean delivery was substantially more common among pregnancies complicated by preterm birth (16.39% vs 7.10%), previous preterm birth (4.39% vs 3.07%), pre-pregnancy diabetes (1.86% vs 0.60%), gestational diabetes (9.27% vs 6.00%), pre-pregnancy hypertension (3.68% vs 1.69%), and gestational hypertension (10.40% vs 6.48%) (all P<.001). Prior cesarean delivery demonstrated the strongest association, occurring in 41.50% of cesarean births compared with 3.05% of vaginal births (Table 1).

Socioeconomic patterns were also observed. Individuals with cesarean delivery were slightly more likely to have attained a bachelor’s degree or higher (34.70% vs 33.12%), whereas those with vaginal delivery were more likely to have less than a high school education (13.30% vs 11.19%) (P<.001). Gestational age differed markedly, with term deliveries accounting for 83.61% of cesarean births compared with 92.90% of vaginal births (P<.001). Distribution across birth years remained relatively stable over the study period, although modest differences were statistically significant given the large sample size (Table 1).

From 2014 to 2024, vaginal delivery remained the predominant mode of delivery, accounting for approximately two-thirds of all births throughout the study period (Figure 1). The proportion of cesarean deliveries was relatively stable, comprising approximately one-third of deliveries annually, with minimal year-to-year variation. A modest increase in the proportion of cesarean deliveries was observed after 2020, accompanied by a corresponding slight decline in vaginal deliveries; however, overall patterns remained largely consistent across the decade (Figure 1).

**Figure 1.**
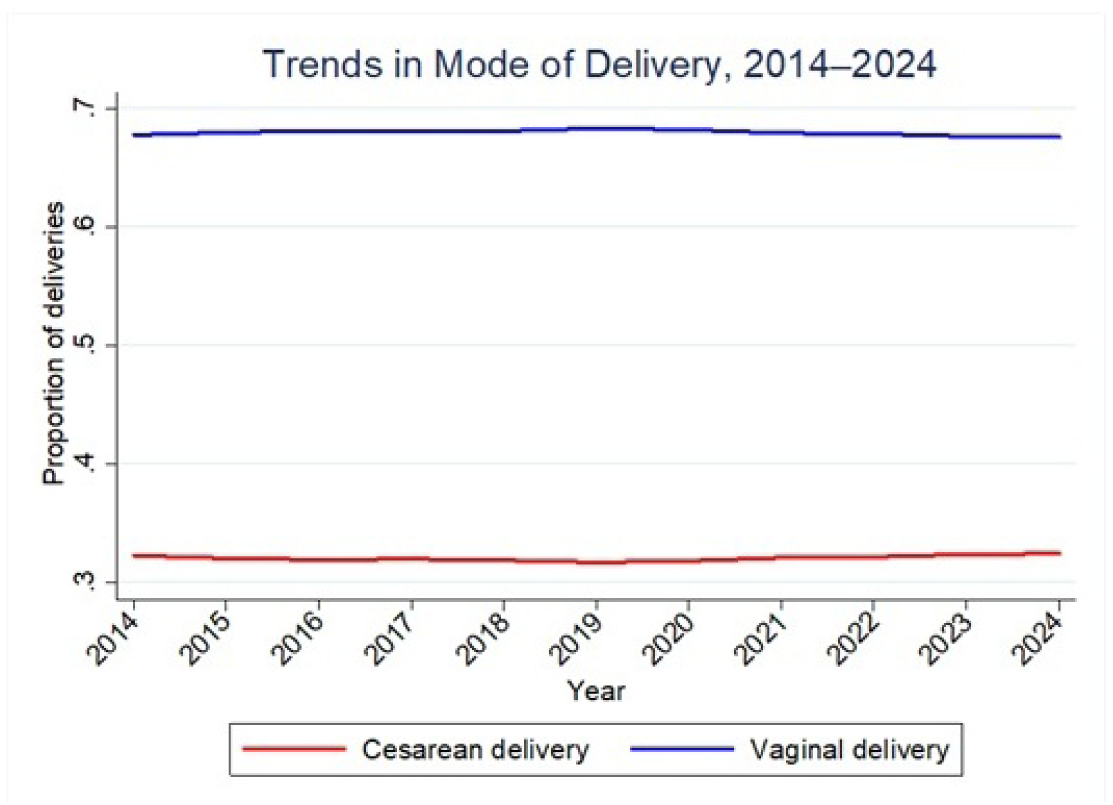
Trends in Cesarean Delivery.

#### Adjusted Independent Associations of Race, Ethnicity, and Insurance Status with Cesarean Delivery

After multivariable adjustment, race/ethnicity and insurance status were independently associated with cesarean delivery (Table 2). Compared with Non-Hispanic White individuals, Non-Hispanic Black individuals had higher odds of cesarean delivery (OR, 1.22; 95% CI, 1.22–1.23), whereas Hispanic individuals had slightly higher odds (OR,1.07; 95% CI, 1.06–1.07). Individuals identifying as Non-Hispanic AIAN or Asian also had lower odds of cesarean delivery. With respect to insurance coverage, privately insured individuals had higher odds of cesarean delivery compared with uninsured women (OR, 1.59; 95% CI, 1.58–1.60). Medicaid beneficiaries also had substantially higher odds compared to uninsured women (OR, 139; 95% CI, 1.38–1.40). These associations persisted after adjustment for maternal age, socioeconomic factors, and pregnancy-related clinical risk factors, including hypertensive disorders and prior cesarean delivery.

**Table 2.** Adjusted Odds Ratios for Cesarean Delivery by Race/Ethnicity and Insurance Status. Odds ratios were estimated using multivariable logistic regression with robust standard errors. The outcome was cesarean delivery (yes vs no). Models were adjusted for maternal age, marital status, educational attainment, pre-pregnancy hypertension, gestational hypertension, prior cesarean delivery, and other clinical covariates. Odds ratios greater than 1 indicate higher odds of cesarean delivery.

| Characteristic | Adjusted OR (95% CI) | P Value |
| --- | --- | --- |
| Race/Ethnicity (reference: Non-Hispanic White) |  |  |
| Non-Hispanic Black | 1.22 (1.22–1.23) | <.001 |
| Non-Hispanic AIAN | 0.83 (0.81–0.84) | <.001 |
| Non-Hispanic Asian | 1.10 (1.09–1.10) | <.001 |
| Non-Hispanic NHOPI | 0.93 (0.91–0.96) | <0.001 |
| Non-Hispanic Multiracial | 0.99 (0.98–1.00) | 0.061 |
| Hispanic (any race) | 1.07 (1.06–1.07) | <.001 |
| Unknown / not stated | 0.97 (0.95–0.98) | 0.001 |
| Insurance status (reference: Uninsured) |  |  |
| Private | 1.59 (1.58–1.60) | <.001 |
| Medicaid | 1.39 (1.38–1.40) | <.001 |
| Other | 1.34 (1.33–1.34) | <.001 |
| Unknown / not stated | 1.35 (1.33–1.37) | <0.001 |

#### Interaction of Race, Ethnicity, and Insurance Status with Cesarean Delivery

Significant effect modification by insurance status was observed in the association between race/ethnicity and cesarean delivery (Table 3). Among uninsured individuals, Non-Hispanic Black (adjusted odds ratio [aOR], 1.83; 95% CI, 1.63–2.06) and Non-Hispanic Asian individuals (aOR, 1.89; 95% CI, 1.70–2.11) had substantially higher odds of cesarean delivery compared with the reference group which were white women with similar insurance. Elevated odds were also observed among uninsured Hispanic individuals (aOR, 1.29; 95% CI, 1.19–1.40) and Non-Hispanic Native Hawaiian or Other Pacific Islander individuals (aOR, 1.68; 95% CI, 1.05–2.70).

**Table 3.**
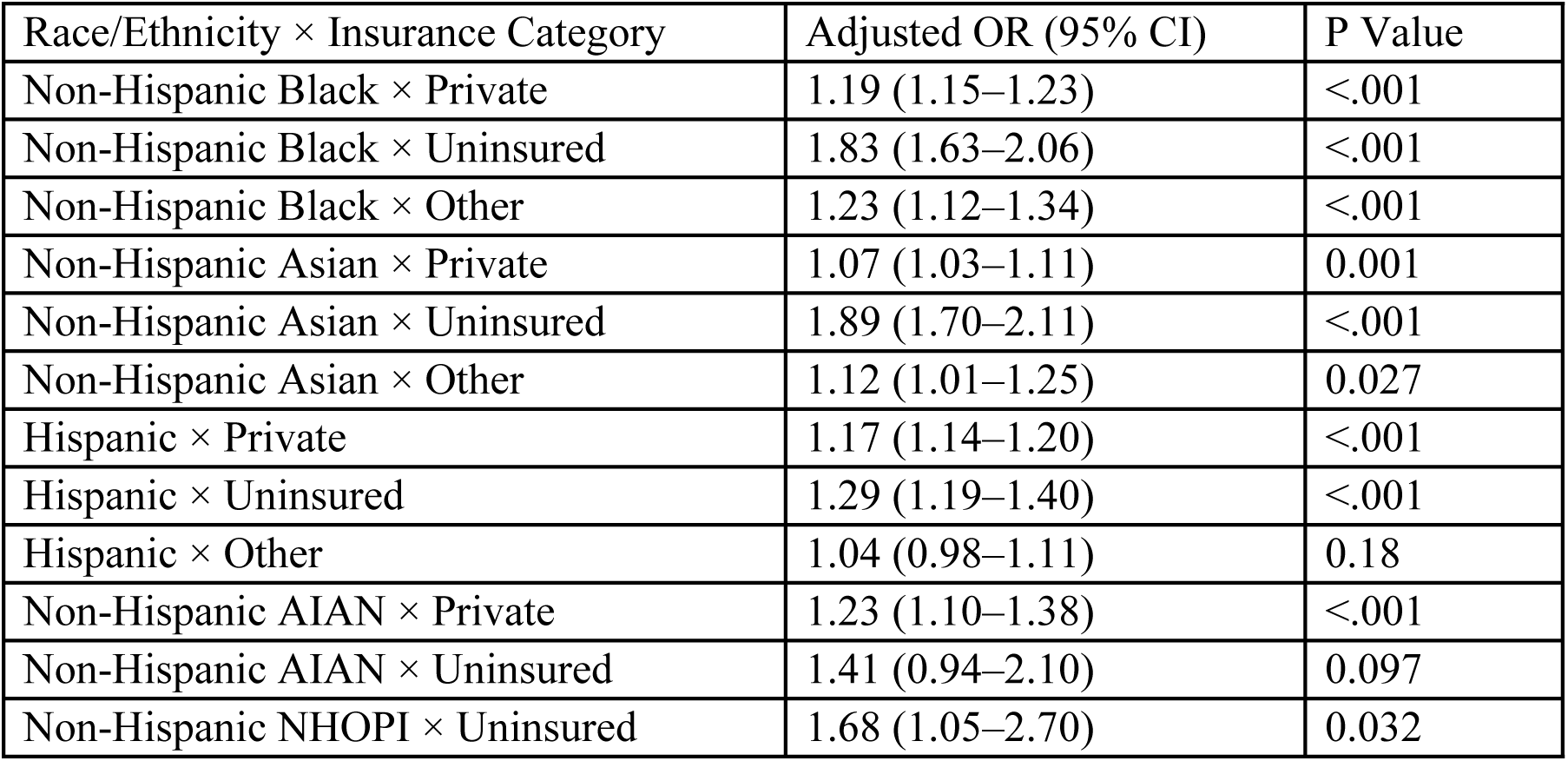
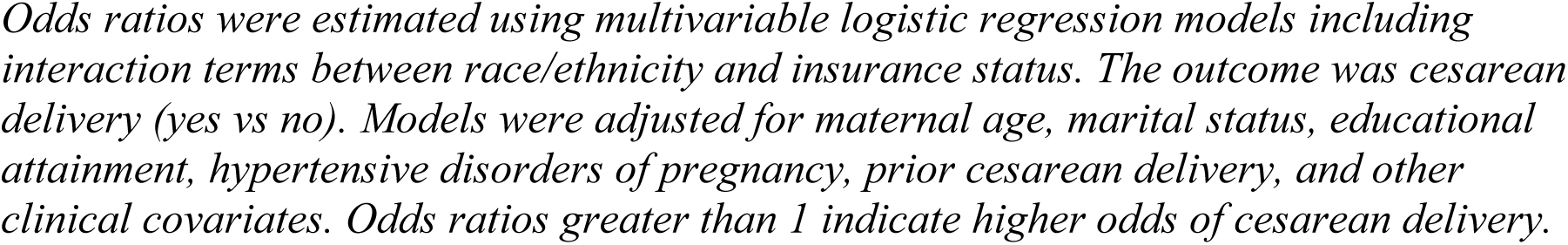
Adjusted Odds Ratios for Cesarean Delivery by Race/Ethnicity and Insurance Type. Odds ratios were estimated using multivariable logistic regression models including interaction terms between race/ethnicity and insurance status. The outcome was cesarean delivery (yes vs no). Models were adjusted for maternal age, marital status, educational attainment, hypertensive disorders of pregnancy, prior cesarean delivery, and other clinical covariates. Odds ratios greater than 1 indicate higher odds of cesarean delivery.

| Race/Ethnicity × Insurance Category | Adjusted OR (95% CI) | P Value |
| --- | --- | --- |
| Non-Hispanic Black × Private | 1.19 (1.15–1.23) | <.001 |
| Non-Hispanic Black × Uninsured | 1.83 (1.63–2.06) | <.001 |
| Non-Hispanic Black × Other | 1.23 (1.12–1.34) | <.001 |
| Non-Hispanic Asian × Private | 1.07 (1.03–1.11) | 0.001 |
| Non-Hispanic Asian × Uninsured | 1.89 (1.70–2.11) | <.001 |
| Non-Hispanic Asian × Other | 1.12 (1.01–1.25) | 0.027 |
| Hispanic × Private | 1.17 (1.14–1.20) | <.001 |
| Hispanic × Uninsured | 1.29 (1.19–1.40) | <.001 |
| Hispanic × Other | 1.04 (0.98–1.11) | 0.18 |
| Non-Hispanic AIAN × Private | 1.23 (1.10–1.38) | <.001 |
| Non-Hispanic AIAN × Uninsured | 1.41 (0.94–2.10) | 0.097 |
| Non-Hispanic NHOPI × Uninsured | 1.68 (1.05–2.70) | 0.032 |

Among privately insured individuals, higher odds of cesarean delivery persisted for Non-Hispanic Black (aOR, 1.19; 95% CI, 1.15–1.23), Hispanic (aOR, 1.17; 95% CI, 1.14–1.20), Non-Hispanic Asian (aOR, 1.07; 95% CI, 1.03–1.11), and Non-Hispanic American Indian or Alaska Native individuals (aOR, 1.23; 95% CI, 1.10–1.38). Associations for Hispanic individuals with other insurance types were not statistically significant (aOR, 1.04; 95% CI, 0.98–1.11).

#### Adjusted Predicted Probabilities of Cesarean Delivery by Race, Ethnicity, and Insurance

Predicted probabilities of cesarean delivery differed meaningfully across racial and ethnic groups and insurance categories (Table 4). The highest probability was observed among privately insured Non-Hispanic Black patients (0.495; 95% CI, 0.490–0.500), whereas uninsured Non-Hispanic White patients had one of the lowest probabilities (0.346; 95% CI, 0.335–0.356).

**Table 4.**
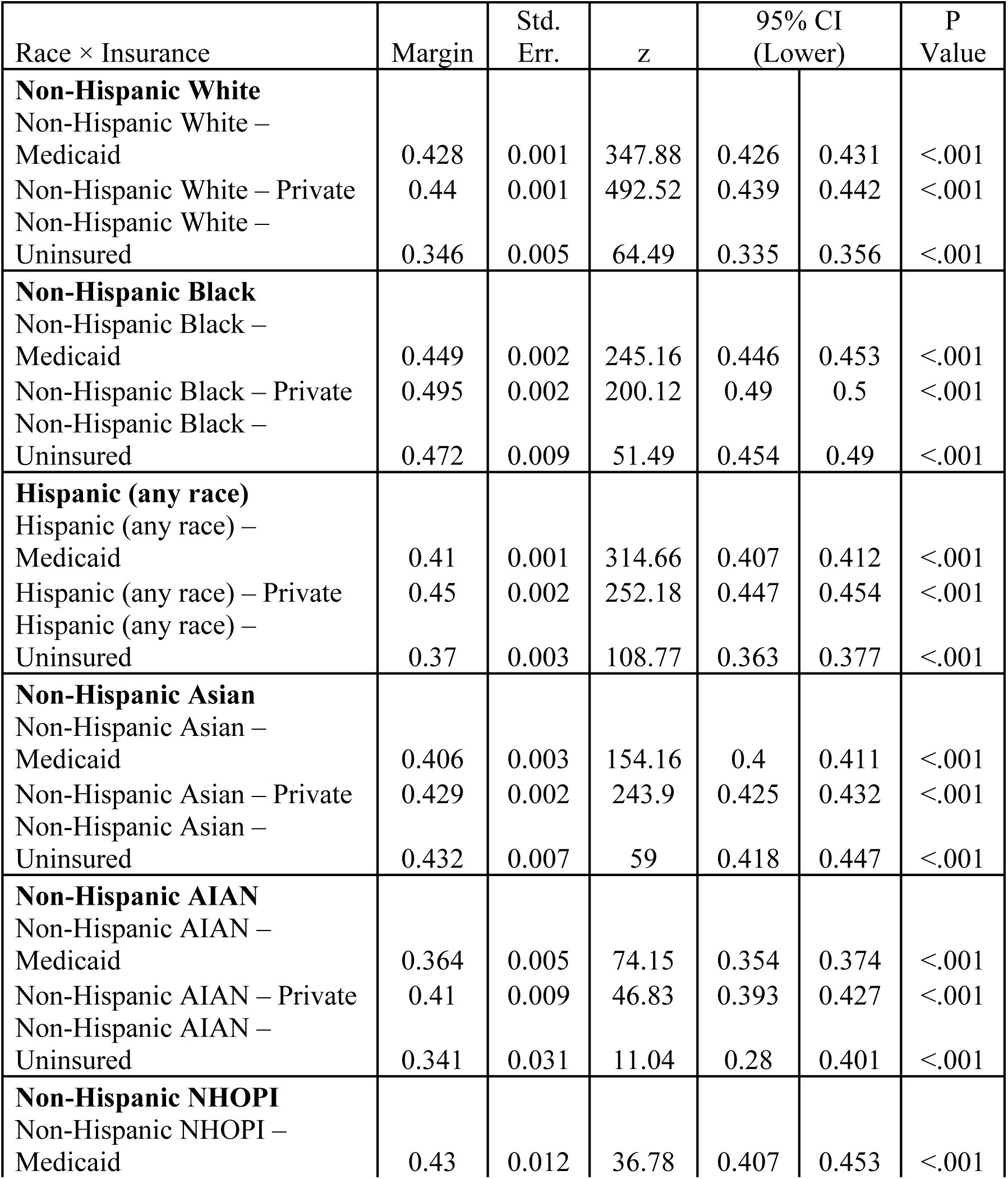
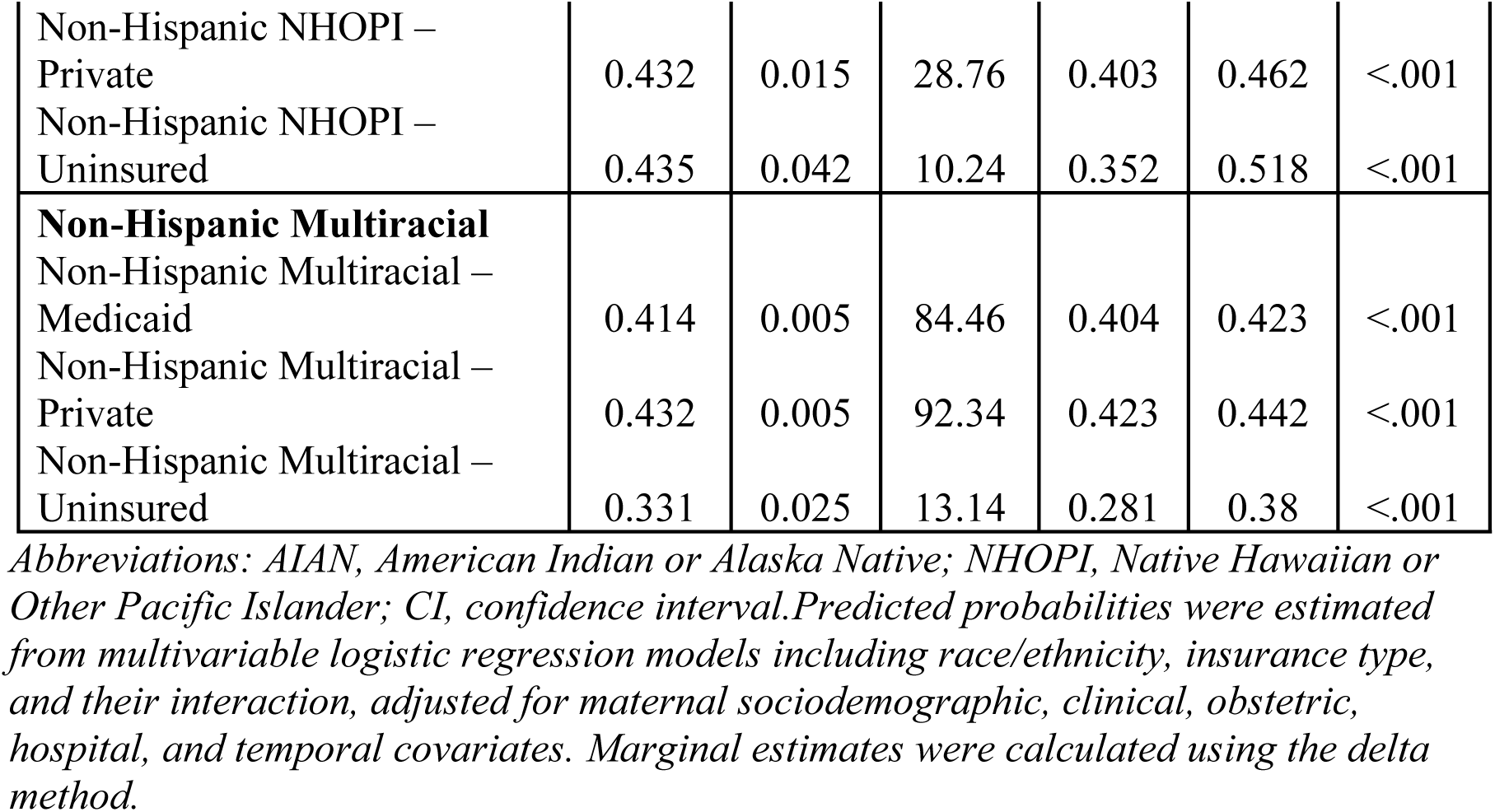
Adjusted Predicted Probabilities of Cesarean Delivery by Race/Ethnicity and Insurance Type, United States.

| Race × Insurance | Margin | Std. Err. | z | 95% CI<br>(Lower) |  | P Value |
| --- | --- | --- | --- | --- | --- | --- |
| **Non-Hispanic White** |  |  |  |  |  |  |
| Non-Hispanic White – Medicaid | 0.428 | 0.001 | 347.88 | 0.426 | 0.431 | <.001 |
| Non-Hispanic White – Private | 0.44 | 0.001 | 492.52 | 0.439 | 0.442 | <.001 |
| Non-Hispanic White – Uninsured | 0.346 | 0.005 | 64.49 | 0.335 | 0.356 | <.001 |
| **Non-Hispanic Black** |  |  |  |  |  |  |
| Non-Hispanic Black – Medicaid | 0.449 | 0.002 | 245.16 | 0.446 | 0.453 | <.001 |
| Non-Hispanic Black – Private | 0.495 | 0.002 | 200.12 | 0.49 | 0.5 | <.001 |
| Non-Hispanic Black – Uninsured | 0.472 | 0.009 | 51.49 | 0.454 | 0.49 | <.001 |
| **Hispanic (any race)** |  |  |  |  |  |  |
| Hispanic (any race) – Medicaid | 0.41 | 0.001 | 314.66 | 0.407 | 0.412 | <.001 |
| Hispanic (any race) – Private | 0.45 | 0.002 | 252.18 | 0.447 | 0.454 | <.001 |
| Hispanic (any race) – Uninsured | 0.37 | 0.003 | 108.77 | 0.363 | 0.377 | <.001 |
| **Non-Hispanic Asian** |  |  |  |  |  |  |
| Non-Hispanic Asian – Medicaid | 0.406 | 0.003 | 154.16 | 0.4 | 0.411 | <.001 |
| Non-Hispanic Asian – Private | 0.429 | 0.002 | 243.9 | 0.425 | 0.432 | <.001 |
| Non-Hispanic Asian – Uninsured | 0.432 | 0.007 | 59 | 0.418 | 0.447 | <.001 |
| **Non-Hispanic AIAN** |  |  |  |  |  |  |
| Non-Hispanic AIAN – Medicaid | 0.364 | 0.005 | 74.15 | 0.354 | 0.374 | <.001 |
| Non-Hispanic AIAN – Private | 0.41 | 0.009 | 46.83 | 0.393 | 0.427 | <.001 |
| Non-Hispanic AIAN – Uninsured | 0.341 | 0.031 | 11.04 | 0.28 | 0.401 | <.001 |
| **Non-Hispanic NHOPI** |  |  |  |  |  |  |
| Non-Hispanic NHOPI – Medicaid | 0.43 | 0.012 | 36.78 | 0.407 | 0.453 | <.001 |
| Non-Hispanic NHOPI – Private | 0.432 | 0.015 | 28.76 | 0.403 | 0.462 | <.001 |
| Non-Hispanic NHOPI – Uninsured | 0.435 | 0.042 | 10.24 | 0.352 | 0.518 | <.001 |
| **Non-Hispanic Multiracial** |  |  |  |  |  |  |
| Non-Hispanic Multiracial – Medicaid | 0.414 | 0.005 | 84.46 | 0.404 | 0.423 | <.001 |
| Non-Hispanic Multiracial – Private | 0.432 | 0.005 | 92.34 | 0.423 | 0.442 | <.001 |
| Non-Hispanic Multiracial – Uninsured | 0.331 | 0.025 | 13.14 | 0.281 | 0.38 | <.001 |

*Abbreviations: AIAN, American Indian or Alaska Native; NHOPI, Native Hawaiian or Other Pacific Islander; CI, confidence interval. Predicted probabilities were estimated from multivariable logistic regression models including race/ethnicity, insurance type, and their interaction, adjusted for maternal sociodemographic, clinical, obstetric, hospital, and temporal covariates. Marginal estimates were calculated using the delta method.*

Among Hispanic patients, probabilities ranged from 0.370 (95% CI, 0.363–0.377) among the uninsured to 0.450 (95% CI, 0.447–0.454) among the privately insured. Similar patterns were observed among Non-Hispanic Asian patients, with probabilities of 0.406 (95% CI, 0.400–0.411) for Medicaid and 0.429 (95% CI, 0.425–0.432) for private insurance (Table 4).

Non-Hispanic American Indian or Alaska Native patients had lower probabilities overall, particularly among the uninsured (0.341; 95% CI, 0.280–0.401), as did uninsured Multiracial patients (0.331; 95% CI, 0.281–0.380). Within nearly all insurance categories, Non-Hispanic Black patients had the highest predicted probabilities (Table 4).

#### Adjusted Risk Differences in Cesarean Delivery by Race and Ethnicity Within Insurance Groups

Adjusted absolute differences in cesarean delivery varied across racial and ethnic groups within insurance categories (Table 5). The largest disparity was observed among uninsured Non-Hispanic Black women, who had a 12.3–percentage point higher risk of cesarean delivery compared with uninsured Non-Hispanic White women (risk difference [RD], 0.123; 95% CI, 0.103–0.144). Elevated risks were also observed among uninsured Hispanic women (RD, 0.084; 95% CI, 0.067–0.102) and uninsured Non-Hispanic American Indian or Alaska Native women (RD, 0.087; 95% CI, 0.005–0.170).

**Table 5.**
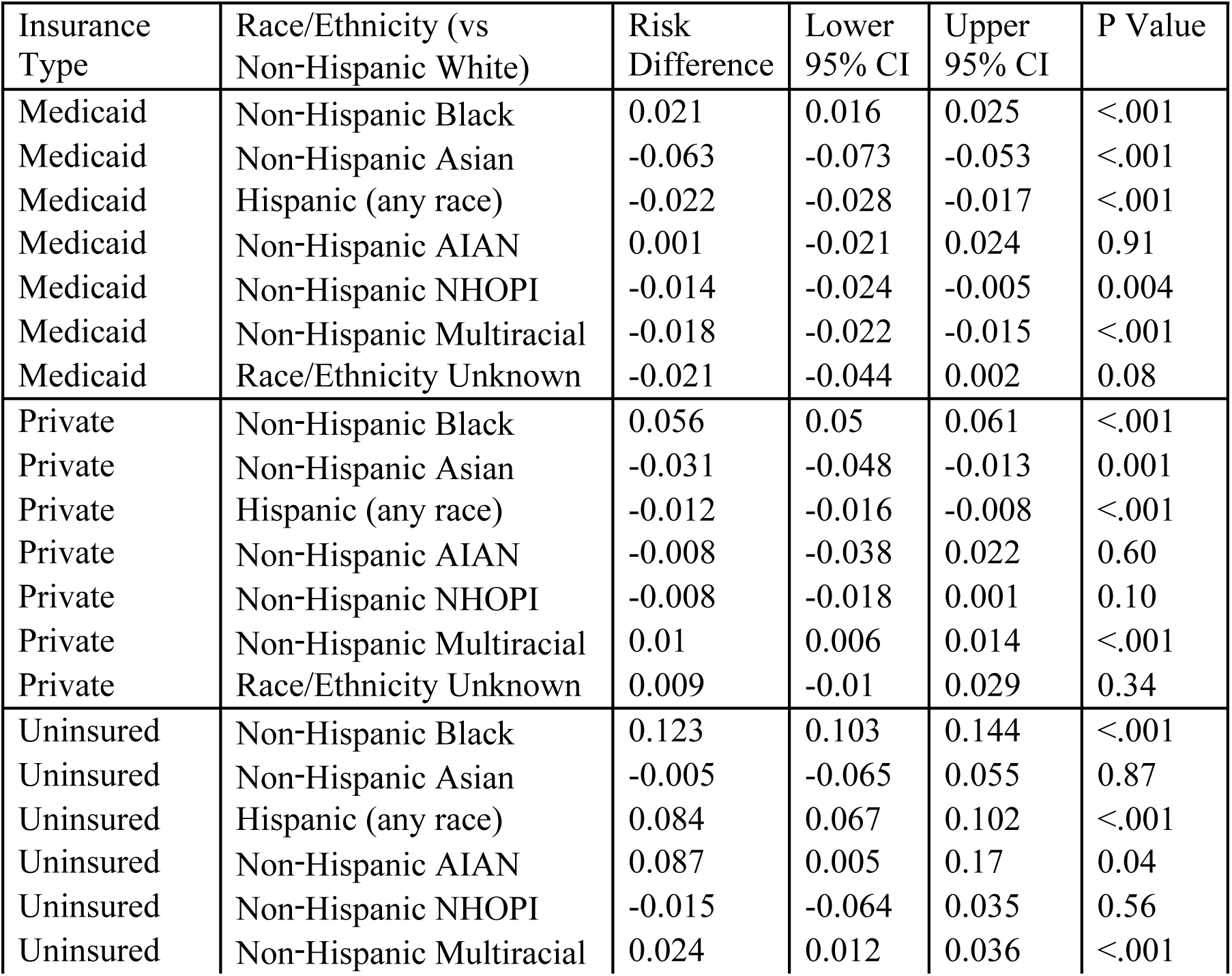
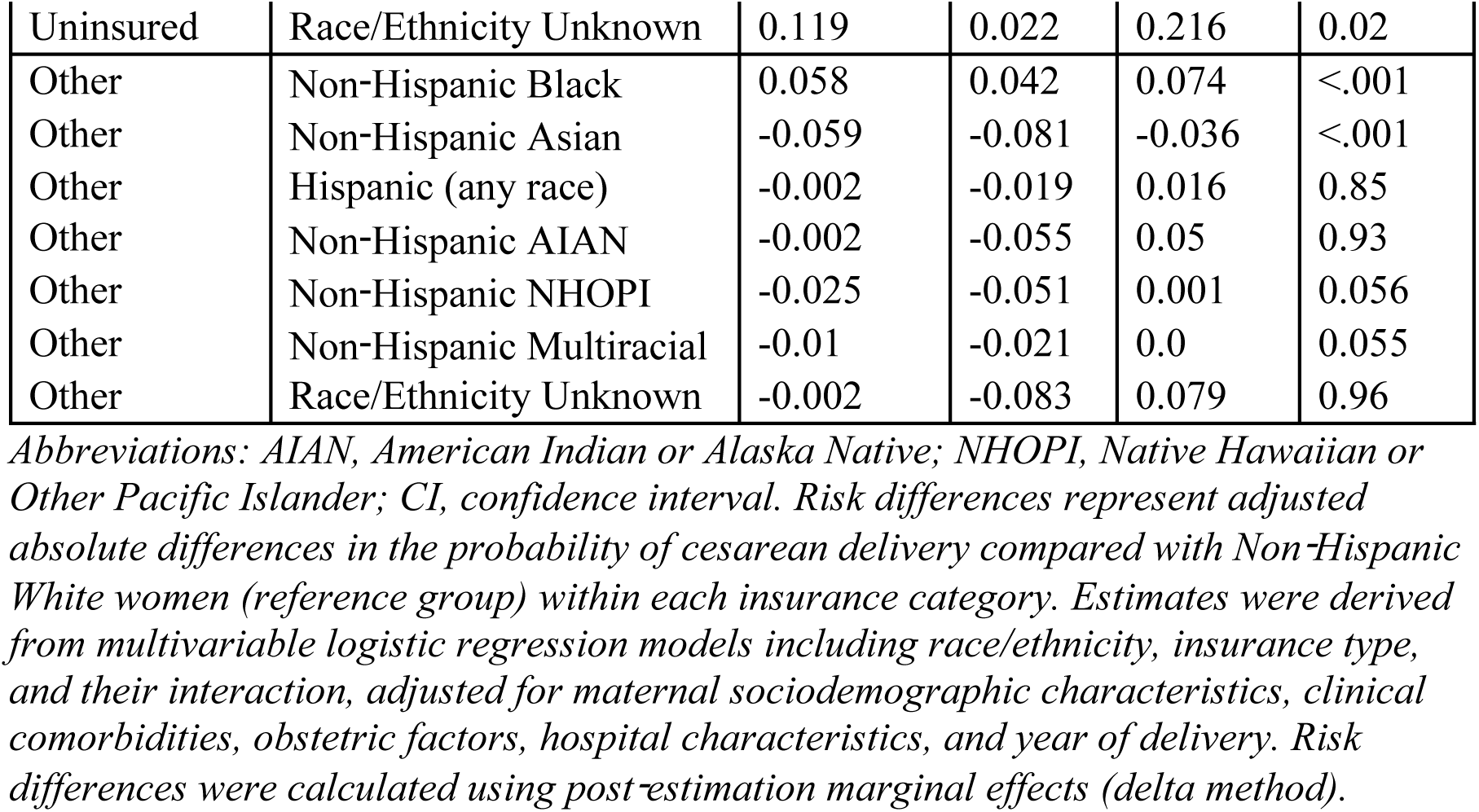
Adjusted Risk Differences in Cesarean Delivery by Race/Ethnicity Compared with Non-Hispanic White Women, Stratified by Insurance Type, United States.

Among privately insured women, Non-Hispanic Black women continued to demonstrate higher risk (RD, 0.056; 95% CI, 0.050–0.061), whereas Hispanic women had slightly lower risk relative to Non-Hispanic White women (RD, −0.012; 95% CI, −0.016 to −0.008). Non-Hispanic Asian women had consistently lower risks across Medicaid (RD, −0.063; 95% CI, −0.073 to −0.053) and private insurance (RD, −0.031; 95% CI, −0.048 to −0.013).

Risk differences for Native Hawaiian or Other Pacific Islander and American Indian or Alaska Native women varied across insurance categories, with several estimates crossing the null, indicating greater statistical uncertainty. Overall, absolute disparities were most pronounced among uninsured populations (Table 5).

## Discussion

In this large, population-based study of U.S. births from 2014 to 2024, cesarean delivery rates remained relatively stable nationally, yet substantial racial and ethnic disparities persisted. Insurance coverage modified these differences but did not eliminate them, suggesting that aggregate stability in cesarean use may conceal important inequities in obstetric care. Together, these findings underscore the complex and intersecting roles of race, ethnicity, and insurance in shaping delivery practices in the United States.

### National Trends in Cesarean Delivery, 2014–2024

Despite substantial changes in obstetric practice, health policy, and population risk profiles over the past decade, cesarean delivery rates in the United States remained largely stable from 2014 to 2024, consistently accounting for approximately one-third of all births [30–32]. This pattern suggests that national efforts to reduce unnecessary cesarean deliveries may have been counterbalanced by competing forces, including rising maternal age, increasing prevalence of comorbidities, greater obstetric complexity, and evolving clinical risk tolerance [6, 33–35]. Similar stability in cesarean delivery rates has been reported in prior national analyses of U.S. birth data, including studies using Vital Statistics that documented a plateau in cesarean use following earlier declines in the mid-2010s [32, 36, 37]. While the relative stability of overall cesarean rates may suggest limited aggregate change, it may also mask substantial heterogeneity across population subgroups [38,39]. Accordingly, examining the independent and interactive roles of insurance coverage and race and ethnicity remains critically important, as stable national trends can coexist with persistent and potentially widening inequities in obstetric care delivery and decision-making.

### Independent and Interactive Effects of Race/Ethnicity and Insurance

Consistent with prior work, Non-Hispanic Black women experienced higher odds and absolute risks of cesarean delivery compared with Non-Hispanic White women across all insurance categories [8,9]. In contrast, American Indian or Alaska Native women generally had lower odds, whereas Hispanic and Non-Hispanic Asian women exhibited more modest and insurance-dependent differences relative to Non-Hispanic White women [11,14]. Insurance status was independently associated with cesarean delivery, with higher rates observed among privately insured women and lower rates among uninsured women relative to Medicaid beneficiaries.

Importantly, our interaction analyses demonstrated that insurance status modified racial and ethnic disparities in cesarean delivery. Disparities were most pronounced among uninsured women, particularly for Non-Hispanic Black women, who exhibited the highest predicted probabilities and largest absolute risk differences compared with Non-Hispanic White women. These findings suggest that insurance coverage does not merely operate as an independent determinant of cesarean delivery but interacts with race and ethnicity to shape patterns of obstetric intervention [40–42].

### Potential Explanations for Observed Disparities

Several mechanisms may underlie the observed independent and interactive effects. Structural racism and implicit bias within healthcare systems may influence clinical decision-making, communication, and thresholds for surgical intervention. Differences in hospital quality, resource availability, and practice patterns may further contribute to variation in cesarean delivery by race and insurance type [16,24,43–45]. Financial and reimbursement incentives associated with private insurance may promote higher intervention rates, while uninsured women may experience delayed presentation, limited access to prenatal care, or differential management during labor [46–48]. Together, these factors may amplify disparities among marginalized populations, particularly when multiple structural disadvantages intersect.

### Comparison with Prior Literature

Our findings align with a substantial body of literature documenting higher cesarean delivery rates among Non-Hispanic Black women compared with White women, even after accounting for clinical risk [10–12]. Prior studies have also reported higher cesarean rates among privately insured women and lower rates among uninsured women [21,22]. However, many earlier studies relied on older data or did not explicitly examine interaction effects between race/ethnicity and insurance status [9,10]. By using a decade of contemporary national data and quantifying disparities using both relative and absolute measures, our study extends existing evidence and demonstrates that insurance coverage meaningfully modifies racial and ethnic disparities in cesarean delivery.

### Population Health and Policy Implications

From a population health perspective, persistent and modified differences in cesarean delivery have important implications for maternal morbidity, healthcare costs, and long-term reproductive health [49–51]. Policies aimed at reducing unnecessary cesarean delivery should incorporate equity-focused strategies that address both racial bias and insurance-related differences in access and care delivery. Quality improvement initiatives, standardized labor management protocols, and accountability metrics stratified by race and insurance may help reduce unwarranted variation. Expanding access to comprehensive prenatal and intrapartum care for uninsured and underinsured populations may also mitigate observed disparities.

### Strengths and Limitations

Key strengths of this study include the use of a large, nationally representative dataset spanning a decade, the ability to examine detailed race and ethnicity categories, and the application of interaction-based analyses with both relative and absolute effect measures. Limitations include the observational design, which precludes causal inference, potential misclassification of race/ethnicity and insurance status on birth certificates, and the absence of detailed clinical information such as labor progression, patient preferences, and provider-level factors. Insurance status at delivery may not fully capture coverage continuity throughout pregnancy.

### Future Research Directions

Future studies should explore hospital- and provider-level contributors to observed disparities, including institutional policies, staffing models, and regional variation. Qualitative research examining patient and provider perspectives may further elucidate mechanisms driving differential cesarean use. Longitudinal analyses assessing changes over time and evaluating the impact of policy interventions aimed at improving equity in obstetric care are also warranted.

### Conclusion

In conclusion, despite relatively stable national cesarean delivery rates from 2014 to 2024, substantial racial and ethnic disparities persist and are significantly modified by insurance coverage. These findings suggest that aggregate stability in cesarean use may mask important inequities in obstetric care and highlight the potential contribution of structural and coverage-related factors to these differences.

## Data Availability

The data used in this study are publicly available from the National Center of Health Statistics (NCHS). Data can be assessed at: https://www.cdc.gov/nchs/index.html For additional information regarding data access, researchers may contact the NCHS National Vital Statistics System at: The CDC serves as the non-author institutional point of contact and maintains long term data availability.

## Access to Data

Dr. Akinyemi had full access to all the data in the study and took responsibility for the integrity of the data and the accuracy of the data analysis.

